# Estimation of COVID-19 outbreak size in Italy based on international case exportations

**DOI:** 10.1101/2020.03.02.20030049

**Authors:** Ashleigh R. Tuite, Victoria Ng, Erin Rees, David Fisman

## Abstract

Italy is currently experiencing an epidemic of COVID-19 which emerged in the Lombardy region ^1^. During the interval between February 25-29, 2020, we identified 46 cases of COVID-19 reported in 21 countries in Europe, Africa, North America, and South America which were either in individuals with recent travel from Italy, or who had presumed infection by a traveler from Italy ^2^. In six cases, in four of the affected countries (Switzerland, France, Austria, Croatia), land travel was a likely route of introduction, or was documented to have been the route of introduction^2^.

---

We used air travel volume between Italian cities and cities in other countries as an index of connectedness, using data available from the International Air Transport Association (IATA) for February 2015 (2.61 million total departing international air passengers from Italy). We used the methods of Fraser et al^3^ to estimate the size of the underlying epidemic in Italy necessary in order for these cases to be observed with a reasonable probability. To estimate the time at risk of COVID-19 exposure for travelers departing Italy, we obtained data from the United Nations World Tourism Organization (UNWTO) for the proportion of international travelers that are non-residents of Italy (63%)^4^ and the average length of stay of tourists to Italy (3.4 days)^5^, and assumed the Italian epidemic began one month prior to February 29, 2020 ^6^.

We also performed sensitivity analyses in which we included outbound travel to all countries regardless of reported case importations, inflated travel volumes by 35%, to account for the relative increase in flight numbers from 2015-2019, and excluded cases in bordering countries and which were documented to have been introduced by overland travel.

When all cases were considered we estimated a true outbreak size of 3971 cases (95% CI 2907-5297), as compared to a reported case count of 1128 on February 29, 2020, suggesting non-identification of 72% (61-79%) of cases. In sensitivity analyses, outbreak sizes varied from 1552 to 4533 cases (implying non-identification of 27-75% of cases) (**Table**).

We recently used similar methods to estimate a much larger epidemic size in Iran, with a far greater degree of under-reporting, based on many fewer exported cases. The reason for this difference relates to the relatively high volume of travel from Italy, relative to Iran^7^. In summary, we suggest that the numerous COVID-19 case exportations from Italy in recent days suggest an epidemic that is larger than official case counts suggest, and which is approximately on a par with that currently occurring in South Korea, which reports 3526 cases (and fewer deaths) as of February 29, 2020^2^.

**Table:** Estimated COVID-19 Outbreak Size, Italy, February 29, 2020.

**Table: Estimated COVID-19 Outbreak Size, Italy, February 29, 2020.**
| **Scenario** | **Estimate** | **95% CI** | **% Unreported\*** | **Unreported Range (%)\*** |
| --- | --- | --- | --- | --- |
| All cases | 3971 | 2907-5297 | 0.72 | 61-79 |
| All cases (adjusted to 2019) | 2937 | 2150-3917 | 0.62 | 48-71 |
| Exclude bordering countries and overland travel | 4533 | 3238-6172 | 0.75 | 65-82 |
| Exclude bordering countries and overland travel (adjusted to 2019) | 3352 | 2395-4564 | 0.66 | 53-75 |
| Include travel to all countries | 2099 | 1500-2859 | 0.46 | 25-60 |
| Include travel to all countries (adjusted to 2019) | 1552 | 1109-2114 | 0.27 | 0-47 |

\*Based on reported case count of 1128.

## Data Availability

IATA data are proprietary; we are able to provide a table with travel volumes upon request. Other data are publicly available. R code available upon request.

## References

1. Day M. Covid-19: Italy confirms 11 deaths as cases spread from north. BMJ 2020;368:m757.

2. BNO News. Tracking coronavirus: Map, data and timeline. Available via the Internet at https://bnonews.com/index.php/2020/02/the-latest-coronavirus-cases/. Last accessed February 29, 2020. 2020.

3. Fraser C, Donnelly CA, Cauchemez S, et al. Pandemic potential of a strain of influenza A (H1N1): early findings. Science 2009;324:1557–61.

4. United Nations World Tourism Organization (UNWTO) : Tourism Statistics: Italy: Country-specific: Basic indicators (Compendium) 2014 - 2018 (12.2019). Aviailable via the Internet at https://www.e-unwto.org/doi/abs/10.5555/unwtotfb0380010020142018201912. Last accessed February 29, 2020.

5. United Nations, Encyclopedia of the Nations: Average length of stay of visitors - Tourism indicators - UNCTAD Handbook of Statistics - Country Comparison. Available via the Internet at https://www.nationsencyclopedia.com/WorldStats/UNCTAD-average-length-stay-visitors.html. Last accessed February 29, 2020.

6. Goes de Jesus J, Sacchi C, Claro I, et al. First report of COVID-19 in South America; available via the Internet at http://virological.org/t/first-report-of-covid-19-in-south-america/409. Last accessed February 29, 2020. 2020.

7. Tuite AR, Bogoch I, Sherbo R, Watts A, Fisman DN, Khan K. Estimation of COVID-2019 burden and potential for international dissemination of infection from Iran. medRxiv 2020.

